# Self-other voice confusion in patients with auditory-verbal hallucinations and nonclinical hallucination proneness

**DOI:** 10.1101/2025.10.06.25337403

**Authors:** Jakša Vukojević, Luka Jelić, Kieren McCormack, Jelena Sušac, Ivan Muselimović, Mihovil Bagarić, Petrana Brečić, Volker Dellwo, Mario Cifrek, Aleksandar Savić, Pavo Orepić

**Affiliations:** University Psychiatric Hospital Vrapče, Zagreb, Croatia; University of Zagreb, Faculty of Electrical Engineering and Computing, Zagreb, Croatia; Ericsson Nikola Tesla, Zagreb, Croatia; University of Zurich, Department of Computational Linguistics, Zurich, Switzerland; School of Medicine, University of Zagreb, Zagreb, Croatia; University of Zurich, Linguistic Research Infrastructure, Zurich, Switzerland

**Keywords:** auditory-verbal hallucinations, schizophrenia, self-other voice discrimination, self-recognition, hallucination proneness, cognitive biomarker

## Abstract

**Background and Hypothesis:** Auditory-verbal hallucinations (AVH), hearing voices without external speakers, are a core symptom of schizophrenia. A prominent account proposes that AVH reflect failures to recognize self-generated speech. Similar effects in hallucination-prone individuals suggest that these mechanisms span a continuum from subclinical to clinical manifestations. We used a self-other voice discrimination (SOVD) task, previously identified as a potential biomarker of self-disturbance, to assess self-recognition deficits in patients with (AVH+) and without (AVH−) a history of AVH, as well as in healthy individuals.

**Study Design:** 41 schizophrenia patients (23 AVH+, 18 AVH−) and 40 healthy controls completed the SOVD task. In a follow-up experiment, 26 additional healthy participants performed a familiar-other voice discrimination task identical to SOVD, but without the self-voice. In patients, performance was tested in relation to symptom severity (PANSS), and in controls to hallucination proneness scores.

**Study Results:** SOVD performance was selectively impaired in AVH+ patients, while AVH− patients did not substantially differ from controls. Across groups, higher PANSS and hallucination proneness scores were specifically related to reduced self-voice recognition, with no impact on other-voice recognition. This effect did not extend to familiar-other voice discrimination.

**Conclusions:** Impaired self-voice recognition is a promising selective marker of AVH occurrence in the acute phase of schizophrenia and extends to hallucination proneness in the general population. These findings support a dimensional view of hallucinations and point to deficits specific to self-voice processing rather than general voice perception. They also highlight SOVD as a promising cognitive biomarker of AVH, with direct implications for early identification and targeted interventions.

## Introduction

Auditory-verbal hallucinations (AVH), or hearing voices in an awake state without outside stimuli, are a very common and highly distressing symptom of schizophrenia^1–3^. With current treatments being ineffective in up to a third of patients^4–6^, there is a pressing need to better understand the underlying mechanisms. According to a prominent account^7–11^, AVH may result from misattributing self-generated thoughts to others, reflecting a deficit in self-monitoring that normally distinguishes internal from external stimuli through feedforward sensorimotor predictions. Experimental evidence supports this account, as patients with AVH show impaired self-recognition across various source-monitoring paradigms (for review, see ^12^). Specifically, patients with and without AVH perform similarly when judging externally generated items (e.g., novel words or other people’s voices), but those with AVH show selective deficits for recognizing self-generated stimuli^12^. Neuroimaging findings parallel this behavioral dissociation: AVH patients display reduced suppression of auditory responses during self-produced speech relative to passive listening^13^, and show altered functional connectivity (mostly in the frontotemporal regions)^11,14–16^, both of which is pointing to failures in distinguishing self-from externally generated signals.

Similar patterns are also observed in healthy individuals with higher proneness to hallucinations, who show reduced sensory attenuation during vocalization^17–19^, and a tendency to misattribute their own voice under perceptual ambiguity^20,21^. Such findings indicate that the cognitive and neural mechanisms underlying AVH are present along a continuum^22,23^, spanning from subclinical experiences to full-blown clinical disorders, with AVH accordingly reported in 10-15% of the general population^24–28^. Studying AVH in healthy individuals is particularly valuable, as it allows researchers to isolate the mechanisms specific to hallucinations while avoiding clinical confounds such as antipsychotic medication, chronic illness, and comorbidities^29–31^. Furthermore, AVHs are frequently found in other psychiatric conditions, such as bipolar disorder, borderline personality disorder, and posttraumatic stress disorder, but also in several neurological conditions, such as epilepsy and migraine, making them an even more prominent psychopathological research phenomenon^32–35^. This notion is one of the reasons why this functional system is one of the key aspects of the Research Domain Criteria (RDoC) initiative from the National Institute of Mental Health (NIMH)^36^, but also similar projects^37^ incorporating transdiagnostic research like ROAMER^38^ and PRISM^39^.

One promising biomarker for assessing self-related deficits is the self-other voice discrimination (SOVD) task^40–42^, which combines voice morphing, bone and air conduction stimuli to measure how individuals distinguish their own voice from others. The task is rapid, objective, and scalable, making it a practical tool for detecting self-related disturbances often missed by standard neuropsychological assessments^43^. Supporting its clinical relevance, a recent study in neurosurgical patients^44^ demonstrated selective impairments in self-voice (versus other-voice) recognition, reflecting difficulties in processing self-related information independent of lesion location or general cognitive and auditory function. In another case study^45^, a patient who developed borderline personality disorder after meningioma resection showed a self-other inversion on the SOVD task, directly mirroring her psychiatric symptoms in both behavior and EEG.

Here, we used the SOVD task to investigate self-voice recognition in schizophrenia patients with and without a history of AVH, as well as in healthy individuals varying in hallucination proneness. We hypothesized that self-voice recognition would be impaired in patients with AVH, compared with both patients without AVH and healthy controls, and that healthy participants with higher hallucination proneness would show similar, though milder, deficits. To determine whether these effects are specific to self-processing rather than general voice recognition or attentional deficits, we included a familiar-other voice discrimination task in a separate sample of healthy participants and expected no association with hallucination proneness.

## Methods

### Participants

This study involved 86 adult participants: 43 patients with schizophrenia (23 AVH+, 20 AVH−) and 43 healthy controls. Two participants from the patient group and three from the control group were excluded from further analysis due to technical errors and incomplete data. This resulted in a total of 81 participants: 41 patients with schizophrenia (23 AVH+) and 40 healthy controls.

Inclusion criteria required a confirmed diagnosis of schizophrenia, established through a structured clinical interview conducted by a psychiatrist in accordance with ICD-10 diagnostic criteria^46^. The main exclusion criterion was that the patients were in an acute phase of schizophrenia at the time of enrollment (e.g., having active hallucinations). Other exclusion criteria comprised: the presence of comorbid psychiatric conditions, including other disorders within the schizophrenia spectrum, but also any current or past substance use disorder, neurological disorders, and severe somatic illnesses. The past presence of auditory verbal hallucinations (AVH) was verified through both clinical interviews and review of patients’ electronic health records. Symptom severity was evaluated using the Positive and Negative Syndrome Scale (PANSS)^47^, which was administered by an experienced psychiatrist. None of the patients exhibited hearing impairments, including hearing loss.

Healthy control participants were recruited from the general population. Eligibility criteria included the absence of self-reported psychiatric and neurological disorders, as well as the lack of hearing impairments. An experienced psychiatrist administered clinical scales before enrollment.

Detailed participant information can be found in **Table 1**.

**Table 1.** Participants’ demographic and clinical details.

|  | **Patients**<br>**(AVH+)** | **Patients**<br>**(AVH-)** | **Healthy**<br>**Controls** | **Between-group**<br>**Comparison (P-value)** |
| --- | --- | --- | --- | --- |
| Age | 33.65 ± 9.31 | 37.00 ± 9.46 | 35.50 ± 9.22 | 0.514 |
| Gender (F, M) | 11, 12 | 6, 12 | 20, 20 | 0.484 |
| Hospitalization | 22 | 17 | - | 0.423 |
| Medication (H, A, AE) | 23, 23, 20 | 17, 17, 14 | - | 0.991 |
| PANSS total | 75.83 ± 18.79 | 72.11 ± 15.38 | - | 0.501 |
| PANSS positive | 16.48 ± 5.24 | 13.56 ± 3.76 | - | 0.053 |
| PANSS negative | 23.35 ± 6.85 | 22.56 ± 5.64 | - | 0.694 |
| PANSS general | 36.00 ± 8.49 | 36.00 ± 8.78 | - | 1 |
| CAPS total | - | - | 4.75 ± 3.54 | - |
| CAPS disturbing | - | - | 7.80 ± 7.65 | - |
| CAPS upsetting | - | - | 8.53 ± 7.57 | - |
| CAPS frequency | - | - | 6.83 ± 6.08 | - |
| PDI total | - | - | 3.77 ± 1.93 | - |
| PDI disturbing | - | - | 7.10 ± 4.63 | - |
| PDI thinking | - | - | 6.65 ± 4.85 | - |
| PDI belief | - | - | 10.50 ± 6.46 | - |

*Mean values with standard deviation are presented where applicable. Hospitalization numbers indicate the number of patients that were hospitalized. The Medication section stands for: H - Hypnotic, A - Anxiolytic, AE - Antiepileptic, and the values indicate the number of patients under these medications.*

In the follow-up experiment, we tested 26 additional healthy controls (6 male, mean age ± s.d.: 25.3 ± 2.9 years old) at the University of Zurich. This was done to 1) replicate the main effect observed in Zagreb with a different experimental setup (see *Procedure*); and 2) to show that the observed effect was specific to self-voice, and not to general voice processing (see *Results*).

All participants gave their informed consent in accordance with European Commission directive 2003/10/EC, Declaration of Helsinki Ethical principles for medical research involving human subjects, and institutional guidelines (University Psychiatric Hospital Vrapče, University of Zagreb Faculty of Electrical Engineering and Computing – Ethics Committee registration number 023-01/23-01/1, Ericsson Nikola Tesla, University of Zurich – registration number 25.05.13).

### Procedure

This study used a Self-Other Voice Discrimination (SOVD) task^42^, which combines voice morphing, air-and bone conduction, and psychophysics to quantify self-voice identification. In the task, participants are asked to indicate whether ambiguous self-other voice morphs more closely resemble their own or someone else’s voice. The six voice morph stimuli were designed in the same way as in our previous work^40,41^, and consisted of 15%, 30%, 45%, 55%, 70%, and 85% of participants’ voices (Self). These were presented ten times each in a randomized order, making a total of sixty trials in one experimental block. Each session had four blocks, two with air-conduction headphones and two with bone-conduction headphones, counterbalanced between participants. To prevent stimulus onset predictability, all blocks had inter-trial intervals that jittered between 1.5 and 2 seconds. Participants’ responses were collected by computer mouse, selecting the left button for Self or the right button for Other voice. The participants were not exposed to the sound (recording) of their own voice prior to the experiment. At the end of experiment, each participant filled in Cardiff Anomalous Perceptions Scale (CAPS)^48^ and Peters Delusion Inventory (PDI)^49^ questionnaires, measuring their proneness to hallucinations and delusions, respectively.

In the follow-up experiment in Zurich, participants additionally performed a Familiar-Other Discrimination task, which was structurally identical to SOVD but used a familiar partner’s voice instead of their own. Participants were recruited in gender-matched pairs, each accompanied by a familiar partner (e.g., friend or acquaintance) whose voice was recorded and used as the familiar reference. Both tasks (Self-Other and Familiar-Other) were presented via air conduction, with task order randomized across participants. After the experiment, participants completed the CAPS questionnaire. This design allowed us to test whether the CAPS-performance relationship observed in Zagreb would replicate in an independent cohort, and whether it was specific to self-voice as opposed to general voice perception.

### Stimuli and Materials

The participants’ voices were recorded while vocalizing the phoneme /a/ for 2 to 3 seconds. The recordings were done with a microphone integrated in Huawei CM33 headphones, and each recording was processed with Audacity software to normalize for the average intensity (−12 dBFS), trim to a duration of 500 ms, and clean the background noise. The Huawei CM33 headphones were used as air-conduction medium, while the Aftershokz Sportz Titanium headphones were used as bone-conducting medium. Both headphones were positioned on participant’s head before the start of the experiment, with matched loudness levels adjusted at lower sound intensities to avoid crosstalk. In Zurich, the stimuli were presented with Bose QC24 air-conduction headphones.

Even though in our previous work we used the TANDEM STRAIGHT algorithm^40,41^, here we employed a WORLD Vocoder voice morphing system, which we specifically adapted for clinical use^42^. This system features a graphical interface and fully automated morphing, enabling straightforward use for clinicians without any programming expertise. The present study also provided an opportunity to evaluate its feasibility in a clinical setting. Although using the same voice decomposition process, some acoustic parameters may differ from the TANDEM STRAIGHT-based implementation, potentially leading to perceptual differences. These considerations are further addressed in the *Supplementary Material*. Importantly, to confirm that the findings obtained with the WORLD Vocoder system at the University of Zagreb are consistent with those from TANDEM STRAIGHT morphing, a follow-up experiment at the University of Zurich was conducted with the original TANDEM STRAIGHT procedure (see *Results*).

### Statistical Analysis

The statistical analysis addressed two main questions: 1) whether SOVD performance differed across groups (AVH+, AVH−, controls) and conditions (bone and air conduction), and 2) to assess the relationship between SOVD task performance and clinical measures.

To model SOVD task performance, we applied binomial mixed-effects model with Response (Self vs Other) as dependent variable. Fixed effects predictors included Group (AVH+, AVH−, control), Condition (air vs. bone conduction), and Voice Morph (ranked levels: 15%, 30%, 45%, 55%, 70%, 85%), with a 3-way interaction and a by-participant random intercept.

The relationship between task performance and clinical measures was modeled using linear mixed-effects with task Accuracy (average percentage correct) as the dependent variable. It was modeled separately for patients and controls, with different clinical scores added to the corresponding models – PANSS for patients; CAPS and PDI for controls. Thus, for the patient model, the fixed effects were Group (AVH+, AVH−), Condition (air vs bone), Voice (binary levels: Self, Other), and PANSS (total score), with a 4-way interaction. For the controls model, the fixed effects were Condition (air vs bone), Voice (binary levels: Self, Other), CAPS and PDI total scores, with a 3-way interaction between Condition, Voice, and each scale. Both patient and control models included participant intercepts as random effects.

The model relating CAPS to the Zurich cohort had 3 fixed effects with a 3-way interaction: Task (Self-other, Familiar-other), Dominant Voice (Self/Familiar, Other), and CAPS (score).

Statistical analysis was done in R^50^ using lme4^51^ package for model estimation and lmerTest^52^ for p-values, while the visualization was done using sjPlot^53^ and ggplot2^54^ packages.

## Results

### Task Performance Across Groups

The mixed-effects binomial model showed a significant two-way interaction between Group and Voice Morph, indicating a significant difference across group performances: the AVH+ group had a lower slope than both healthy controls (estimate = −0.112, z = −3.615, p < 0.001) and the AVH− group (estimate = −0.145, z = −3.737, p < 0.001), while the AVH− group did not differ from controls (estimate = 0.034, z = 0.979, p = 0.33) (**Figure 1**). There was no main effect of Group (AVH+ vs AVH−: estimate = 0.575, z = 1.296, p = 0.195; AVH+ vs controls: estimate = 0.467, z = 1.28, p = 0.202; AVH− vs controls: estimate = –0.109, z = –0.27, p = 0.785), and a main effect of Voice Morph (estimate = 0.159, z = 8.97, p < 0.001) showed the expected increase of Self responses with the increase of voice morph levels.

**Figure 1.**
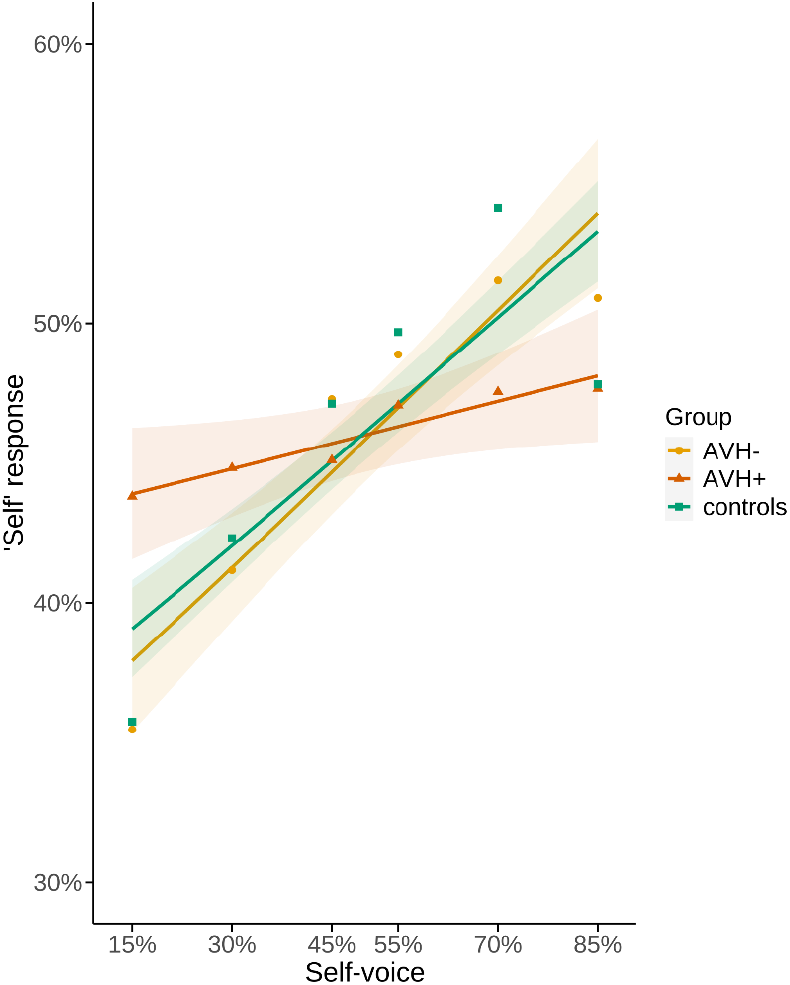
Psychometric curves fitted for the performance in the self-other voice discrimination task across three groups: red for AVH+, yellow for AVH*−*, and green for healthy controls. The x-axis represents the percentage of Self voice in voice morphs, while the y-axis represents the number of Self responses. The shaded area for each curve represents a 95% confidence interval of the local estimates. Task performance dropped specifically in the AVH+ group.

The three-way interaction of Group, Voice Morph, and Condition was not significant (estimate = 0.067, z = 1.55, p = 0.12), implying that the observed group difference was not modulated by air versus bone conduction. A two-way interaction of Condition and Voice Morph (estimate = −0.076, z = −3.05, p = 0.002) revealed a steeper slope for air conduction, confirmed with the corresponding difference in intercepts of Condition (estimate = 0.380, z = 3.90, p < 0.001).

### Task Performance and Clinical Measures

Linear mixed-effects models associating task performance with clinical scores revealed a relationship specific to performance in self-voice trials (**Figure 2**). In the patient model, there was a significant 2-way interaction between Voice and PANSS (estimate = −0.008, t = −4.91, p < 0.001), demonstrating a decrease only in self-voice accuracy as symptom severity increased (**Figure 2a**). Moreover, there was a main effect of Voice (estimate = 0.49, t = 4.04, p < 0.001), demonstrating overall higher accuracy for self-voice, and no main effect of PANSS (estimate = −0.00004, t = −0.02, p = 0.986). The main effect of Group (estimate = −0.698, t = −3.15, p = 0.003), showed, as above, lower performance in AVH+ participants. A 2-way interaction between Group and PANSS (estimate = 0.008, t = 2.86, p = 0.006) indicated a stronger decrease in performance in AVH− group with the increase of PANSS. As exploratory analyses, we ran similar models relating task performance with PANSS subscales, hospitalization, age, and gender, and the results are reported in the *Supplementary Material*.

**Figure 2.**
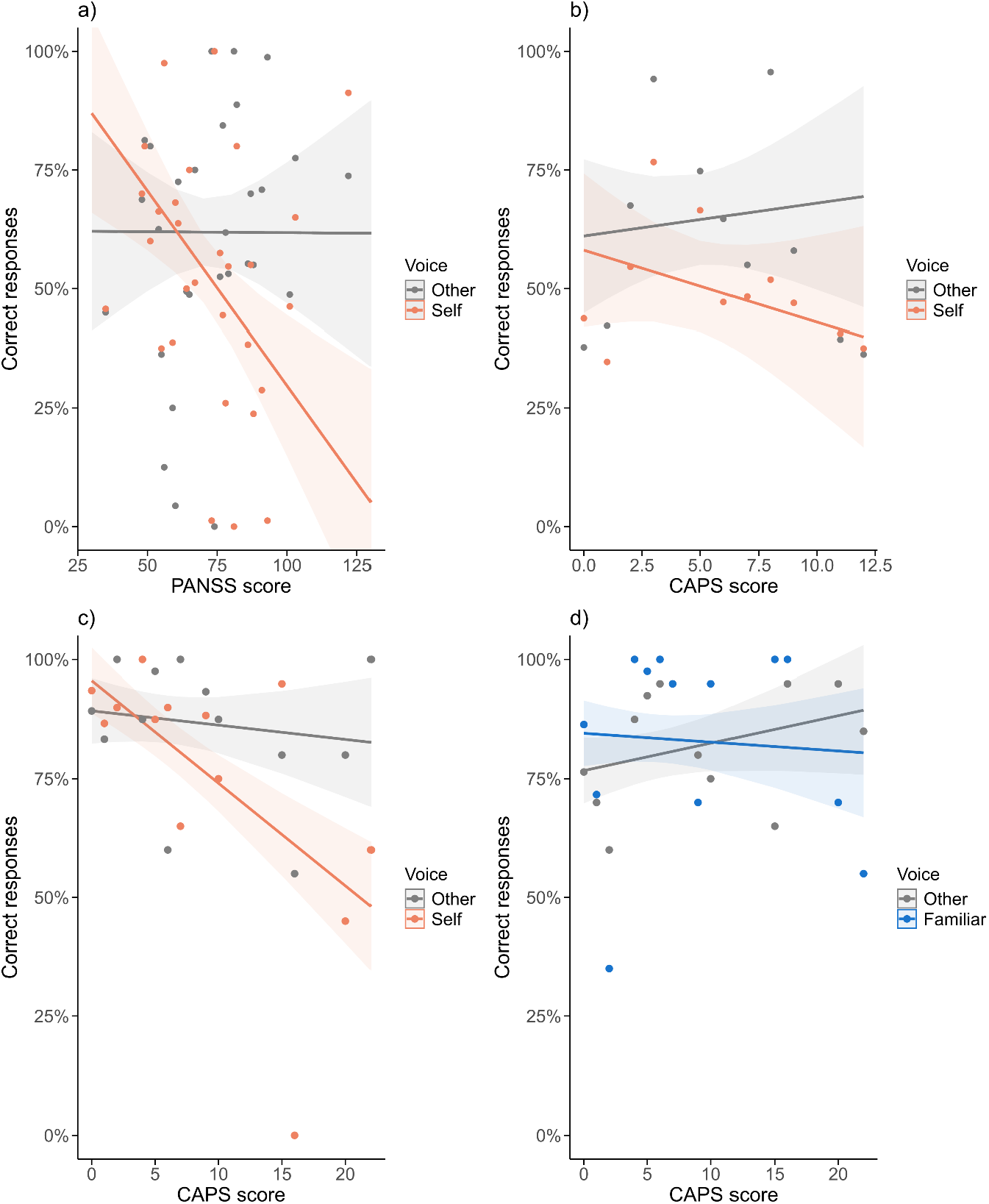
Relationship between SOVD task performance and clinical scores. **a)** Patients: accuracy vs. PANSS, with lines for Self (orange) and Other (grey) voice. **b)** Controls: accuracy vs. CAPS, Self (orange) and Other (grey). **c)** Follow-up controls: accuracy vs. CAPS, Self (orange) and Other (grey). **d)** Follow-up controls: accuracy vs. CAPS, Familiar (blue) and Other (grey) voice. Shaded areas indicate 95% confidence intervals. Task performance drops specifically for self-voice, both in patients and in two control groups.

Mirroring the finding in patients, the model fitted for controls displayed a 2-way interaction between CAPS and Voice (estimate = −0.022, t = −5.08, p < 0.001), showing decrease only in self-voice accuracy with the increase of CAPS scores (**Figure 2b**). There was also a significant 3-way interaction between CAPS, Voice, and Condition (estimate = 0.026, t = 4.27, p < 0.001), showing that this effect was more prominent in air conduction. The main effect of Voice (estimate = −0.083, t = −2.50, p = 0.012) indicated lower accuracy for self-voice, while CAPS alone had no significant main effect (estimate = 0.007, t = 0.48, p = 0.637). There were no significant effects observed for PDI scores (*Supplementary Material*).

To replicate and narrow the CAPS effect specifically to self-voice, as opposed to general voice processing, a follow-up experiment was conducted in which participants performed both a Self-Other and a Familiar-Other Discrimination task. Replicating the initial finding, we found a significant 3-way interaction between CAPS, Task, and Voice (estimate = −0.011, t = −2.27, p = 0.023). When splitting by task, a 2-way interaction between CAPS and Voice was significant only for the Self-Other task (estimate = −0.009, t = −2.58, p = 0.010; **Figure 2c**), again showing a decrease in performance specific to self-voice. The same 2-way interaction was not significant for the Familiar-Other task (estimate = 0.009, t = 1.63, p = 0.111, **Figure 2d)**. There was no main effect of CAPS (estimate = 0.006, t = 1.44, p = 0.157).

Overall, these results demonstrate that both symptom severity in patients and hallucination proneness in controls are related specifically to self-voice recognition, with no effects on the recognition of other familiar or unfamiliar voices.

## Discussion

This study, assessing self-other voice discrimination (SOVD) task performance in schizophrenia patients and healthy controls, has two main findings. First, SOVD was specifically impaired in patients with a history of AVH (AVH+ group), with no statistically significant differences between AVH− patients and healthy participants. Second, clinical symptom severity measured by PANSS and hallucination proneness measured by CAPS were explicitly associated with performance decreases in self-voice trials. The hallucination proneness effect was replicated in an independent cohort of healthy participants and remained specific to self-voice, indicating that it cannot be explained by general deficits in voice perception or attentional demands.

The selective SOVD impairment in AVH+ patients suggests that difficulties in distinguishing self from other voices are not a trait feature of schizophrenia in general but are linked to the occurrence of AVH in the acute phase of the illness. This aligns with prior evidence specifically relating AVH with self-other confusion across various paradigms and modalities^12^. Traditionally, this has been interpreted within the self-monitoring account of AVH^7,8^. According to this framework, motor actions generate sensory predictions that are later compared with actual sensory feedback. When predictions match, feedback is attenuated and attributed to the self; when mismatched, the feedback lacks attenuation and is misattributed to another agent^55–58^. In AVH, it has been proposed that feedforward prediction errors may cause self-generated thoughts to be misattributed, thereby facilitating self-other confusion^7–11^.

Although self-monitoring deficits represent a prominent account of AVH, they cannot fully explain the current findings because the SOVD task does not require active speech production or motor output. Instead, our results point to a broader impairment in self-representation, potentially involving memory processes, or higher-order aspects of self-processing, such as self-identification^59,60^. This perspective aligns with other theoretical accounts of AVH, including models emphasizing deficits in memory processing^61,62^, and overreliance on inherent models of the world^63,64^, in a complementary fashion. For instance, recent theoretical^65^ and empirical^29,66^ work has proposed that self-monitoring processes may help maintain the stability of internal models of self and world, and that disruptions in these processes give rise to self-other confusion. It is thus possible that, in AVH+ patients, self-monitoring deficits accumulate over time (e.g. repeated misattribution of inner speech), resulting in a trait-like self-other confusion, observable across different experimental paradigms, including SOVD. Consistent with this view, we observed SOVD deficits specifically in patients with a history of AVH, supporting the notion of a stable, trait-like alteration rather than a transient, state-dependent effect.

The group differences in SOVD performance suggest that self-other confusion may manifest differently across the psychosis spectrum. AVH+ patients showed a generalized deficit, with flattened psychometric curves indicating a broad inability to distinguish self from other voices. By contrast, AVH− patients and healthy participants performed at a comparable overall level, yet within both groups higher PANSS or CAPS scores were specifically associated with poorer recognition of self-voice, while other-voice performance remained intact. Importantly, this association was replicated in an independent healthy cohort and remained restricted to self-voice, indicating that it is not driven by general voice-perception deficits or task-related attentional demands. Thus, whereas a global SOVD impairment appears characteristic of individuals with a clinical history of AVH, more subtle self-specific vulnerabilities might emerge along the broader psychosis spectrum and modulate performance in a dimensional fashion. In this view, SOVD performance may index a trait-like cognitive marker of vulnerability to AVH, which may become clinically manifest over time due to various additional factors such as stress, cognitive load, or neurobiological changes. Interestingly, the self-voice effect was observed with CAPS but not with PDI, suggesting specificity to hallucination proneness rather than delusional ideation, and pointing to partially dissociable mechanisms within the psychosis continuum. This latter finding should, however, be taken with caution given the limited sample size. Taken together, these findings support dimensional models of psychosis^23–28^, in which mechanisms of self-representation underlie hallucinatory experiences across the population: subtle, self-specific liabilities in nonclinical and AVH− samples may accumulate into global self-processing impairments in AVH+.

Self-voice recognition has previously been used to probe broader deficits in self-processing. Most prior work focused on neurological^67^ and neurosurgical^43^ patients, where SOVD proved useful as a formal neuropsychological tool to quantify post-surgical changes in the sense of self^44^. This approach has also been applied in psychiatry, showing that SOVD can capture altered self-experience in borderline personality disorder^45^ and autistic traits^68^. Building on this foundation, the present study applies SOVD to schizophrenia, directly targeting self-processing mechanisms implicated in AVH. More broadly, these findings underscore the fundamental role of self-processing in human cognition: distinguishing self-generated from external signals is essential for perception, agency, and social interaction, and its disruption can manifest across neurological and psychiatric conditions. From a functional system, or RDoC, perspective^36^, SOVD offers a dimensional, neurobehavioral marker of self-processing that bridges clinical and non-clinical populations. It may also help identify patients with a higher risk of transitioning to psychosis^69^—aligning with the goals of the Accelerating Medicines Partnership– Schizophrenia (AMP-SCZ) program in collaboration with the National Institutes of Health^70^.

## Limitations

A main limitation of our work is the globally weaker performance, present not only in patients but also in healthy controls, likely due to the use of WORLD Vocoder in the SOVD procedure^42^, which replaced the original TANDEM STRAIGHT algorithm for voice morphing. We speculate that the drop in performance may reflect differences in the morphing of higher formant frequencies, which are critical for voice identity recognition^71,72^. This also complicates the interpretation of other acoustics-related effects, such as bone-versus air-conduction performance. The precise differences in acoustics between the two systems and their perceptual effects will be systematically evaluated in our follow-up work. Notably, despite the overall performance drop, the main between-group differences were still observed (Figure 1). It is possible that the TANDEM STRAIGHT morphing, by avoiding putative “floor effects”, would have produced even more pronounced differences in patients. This is supported by our follow-up experiment, in which the relationship between hallucination proneness and self-voice recognition was stronger using TANDEM, compared to WORLD Vocoder morphing (compare Figures 2b and 2c). Finally, a limited sample size further limits the interpretation of finer clinical findings, such as subtle differences regarding the specific PANSS subscales in patients, or differences between CAPS and PDI effects in controls (see *Supplementary Material*).

## Conclusion

Our findings demonstrate that impaired self-other voice discrimination is specifically linked to the occurrence of AVH in the acute phase of the illness and hallucination proneness, supporting the notion that there is an underlying core deficit in self-representation that spans clinical and non-clinical populations. The selective self-voice impairment highlights a trait-like vulnerability for AVH, aligning with dimensional models of psychosis. Future work should aim to disentangle trait versus state mechanisms using longitudinal designs, explore interactions with other cognitive processes implicated in AVH, and map the corresponding neural correlates of these effects.

## Data Availability

All data produced in the present study are available upon reasonable request to the authors

