## Supplementary material for "Self-other voice confusion in patients with auditory-verbal hallucinations and nonclinical hallucination proneness"

### *Voice morphing technologies and SOVD performance difference*

When designing our study, the choice between using one of the two different voice morphing technologies was based on their usability in clinical settings. TANDEM-STRAIGHT[1] morphing application proved to be time consuming and prone to human errors, while the WORLD[2] Vocoder offered its implementation in an easy-to-use Python-based application. Even though previously we used TANDEM-STRAIGHT[3,4], we recently adapted WORLD Vocoder specifically for clinical use[5], and this study offered the first opportunity to evaluate its feasibility in a clinical patient sample. Our new system[5] features a graphical user interface, automated voice morphing, a personalized voice selection tool, and results visualization. This improves task usability, reduces procedure time, and supports a more individualized, patient-centered approach, enabling clinicians to use it without any programming expertise. However, the use of WORLD Vocoder introduced acoustic changes in voice morphs, leading to globally weaker performance. When comparing our healthy control group of 40 participants done with WORLD versus the pilot group of 26 participants done with the TANDEM-STRAIGHT algorithm, the performance differences were evident.

Although TANDEM-STRAIGHT and WORLD Vocoder decompose the voice in the same acoustic components (fundamental frequency, spectral envelope, and aperiodicity), they do it in a different way. While efficient and fast, the WORLD algorithm smooths out the subtle variations in voice pitch, timbre, and higher formants, which are critical for voice identity recognition[6,7], potentially making the morphed voices less distinguishable. On the other hand, TANDEM-STRAIGHT algorithm preserves these imperfect voice details and thus makes the morphs sounding more natural, maintaining the voice signature of the speaker. We therefore speculate that these algorithm differences are the reason behind the flattening effect of the psychometric curve in the main study control group. These differences and their effect on task performance will be formally evaluated in our follow-up work.

### *PANSS subscales*

To further explore the relationship between task performance and clinical measures in patient group, we used linear mixed-effects model with task Accuracy as the dependent variable, and fixed effects of Group (AVH+, AVH-), Voice (binary levels: Self, Other), and PANSS subscales (positive, negative, general), with a 3-way interaction between Group, Voice and each subscale.

As it can be seen from the **Table 1**, a significant 3-way interactions between Group, Voice and PANSS positive and PANSS general subscales were observed. The 2-way interactions show that all PANSS subscales significantly interact with the Voice and Group, except for the negative subscale which does not significantly interact with AVH+ group. Main effects indicate that the task performance drops with higher PANSS general score, while it increases with PANSS positive and PANSS negative scores. As is evident from the significant main effects of all PANSS subscales, the task performance drops with the PANSS general score, while it increases with PANSS positive and PANSS negative. Voice and Group also show significant main effects, with Voice increasing and Group (AVH+) decreasing task accuracy.

**Table 1**. Linear mixed-effects model results: SOVD task accuracy with PANSS subscales.

| **Effect** | **Estimate** | **t value** | **p value** |
|---|---|---|---|
| (Intercept) | 0.577 | 3.353 | 0.00183 ** |
| Group: AVH+ | -0.683 | -3.116 | 0.00349 ** |
| Voice: Self | 0.579 | 6.578 | p < 0.001 *** |
| PANSS: positive | 0.023 | 2.334 | 0.02502 * |
| PANSS: negative | 0.020 | 1.709 | 0.09568 . |
| PANSS: general | -0.020 | -2.788 | 0.00827 ** |
| Group: AVH+ x Voice: Self | -0.129 | -1.151 | 0.24959 |

| | | | |
|---|---|---|---|
| Group: AVH+ x PANSS: positive | -0.031 | -2.043 | 0.04810 * |
| Voice: Self x PANSS: positive | -0.044 | -8.763 | p < 0.001 *** |
| Group: AVH+ x PANSS: negative | -0.002 | -0.179 | 0.85890 |
| Voice: Self x PANSS: negative | -0.055 | -9.432 | p < 0.001 *** |
| Group: AVH+ x PANSS: general | 0.031 | 2.794 | 0.00814 ** |
| Voice: Self x PANSS: general | 0.032 | 8.864 | p < 0.001 *** |
| Group: AVH+ x Voice: Self x PANSS: positive | 0.042 | 5.417 | 6.29e-08 *** |
| Group: AVH+ x Voice: Self x PANSS: negative | 0.010 | 1.424 | 0.15443 |
| Group: AVH+ x Voice: Self x PANSS: general | -0.017 | -2.928 | 0.00342 ** |

These complex and somewhat contradictory results should be taken with caution, as they might result from the relatively small sample size, and a general "floor effect" in AVH+ group (flat average performance), which could have reduced the overall sensitivity and influence the performance across PANSS subscales.

### *PDI score and task accuracy*

The relationship between task performance and clinical measures in controls was investigated using the linear mixed-effects model with task Accuracy as the dependent variable, and fixed effects of Condition (air vs bone), Voice (binary levels: Self, Other), and CAPS and PDI total scores, with a 3-way interaction between Condition, Voice and each scale.

While all the significant main effects have already been described in the main text, here in **Table 2** we can observe that the PDI total score has a tendency for interaction with Voice, suggesting that its role in the model outcome might be similar but less prominent compared to CAPS total score. Since CAPS and PDI scores do correlate[8], this is an expected finding.

**Table 2**. Linear mixed-effects model results: SOVD task accuracy with CAPS and PDI scores for control group.

| Effect | Estimate | t value | p value |
|---|---|---|---|
| (Intercept) | 0.603 | 5.409 | p < 0.001 *** |
| Voice: Self | -0.083 | -2.503 | 0.0123 * |
| CAPS total | 0.007 | 0.475 | 0.6374 |
| Condition: Bone | -0.084 | -2.531 | 0.0114 * |
| PDI total | 0.002 | 0.085 | 0.9325 |
| Voice: Self x CAPS total | -0.022 | -5.083 | p < 0.001 *** |
| Voice: Self x Condition: Bone | -0.010 | -0.223 | 0.8237 |
| CAPS total x Condition: Bone | -0.006 | -1.459 | 0.1445 |

| | | | |
|---|---|---|---|
| Voice: Self x PDI total | 0.015 | 1.903 | 0.0571 . |
| Condition: Bone x PDI total | 0.012 | 1.484 | 0.1378 |
| Voice: Self x CAPS total x Condition: Bone | 0.026 | 4.271 | p < 0.001 *** |
| Voice: Self x Condition: Bone x PDI total | -0.014 | -1.244 | 0.2135 |

### *Hospitalization effects*

For the analysis of hospitalization effect on task performance in patients, we applied linear mixed-effects model with task Accuracy as the dependent variable, and fixed effects of Group (AVH+, AVH-), Voice (binary levels: Self, Other), First Hospitalization (years from first hospitalization), Last Hospitalization (years from first hospitalization), and Number of Hospitalizations, with a 3-way interaction as above.

The results from **Table 3** demonstrate a significant 3-way interactions between the Group, Voice, and the Number of Hospitalizations, and between the Group, Voice, and the Last Hospitalization. There are three 2-way significant interactions: between the Group and Number of Hospitalizations, between the Voice and First Hospitalization, and between Group and Voice. Significant main effects are First Hospitalization and Intercept.

**Table 3**. Linear mixed-effects model results: SOVD task accuracy with hospitalizations for patient group.

| **Effect** | **Estimate** | **t value** | **p value** |
|---|---|---|---|
| (Intercept) | 0.609 | 9.150 | p < 0.001 *** |
| Group: AVH+ | -0.150 | -1.599 | 0.11959 |
| Voice: Self | -0.031 | -1.027 | 0.30461 |
| Last Hospitalization | 0.011 | 1.017 | 0.31692 |
| Last Hospitalization | 0.031 | 1.825 | 0.07726 . |
| Number of Hospitalizations | -0.017 | -1.642 | 0.11038 |
| Group: AVH+ x Voice: Self | 0.194 | 4.504 | p < 0.001 *** |
| Group: AVH+ x First Hospitalization | -0.016 | -1.060 | 0.29688 |
| Voice: Self x First Hospitalization | -0.016 | -3.207 | 0.00135 ** |
| Group: AVH+ x Last Hospitalization | -0.030 | -1.227 | 0.22867 |
| Voice: Self x Last Hospitalization | -0.001 | -0.169 | 0.86619 |
| Group: AVH+ x Number of Hospitalizations | 0.056 | 2.939 | 0.00603 ** |
| Voice: Self x Number of Hospitalizations | 0.006 | 1.301 | 0.19316 |
| Group: AVH+ x Voice: Self x First Hospitalization | 0.011 | 1.629 | 0.10335 |
| Group: AVH+ x Voice: Self x Last Hospitalization | 0.027 | 2.406 | 0.01616 * |
| Group: AVH+ x Voice: Self x Number of Hospitalizations | -0.082 | -9.471 | p < 0.001 *** |

These results suggest that the hospitalization effect might also be self-voice-specific. For all patients, a decrease in task performance is connected to the longer illness duration (years from first hospitalization). The AVH+ group showed an accuracy increase with longer time after last hospitalization, while their performance significantly dropped with the number of hospitalizations.

### *Age and gender effects*

The effects of age and gender for all participant groups were analyzed with the linear mixed-effects model with task Accuracy as the dependent variable, and fixed effects of Group (AVH+, AVH-, controls), Voice (binary levels: Self, Other), Condition (air vs bone), Age (mean = 35.3 years, range = 21–61), with the 3-way interactions between Group, Voice, and Condition. A more complex model with 5-way interactions was disregarded to improve the interpretability.

The results from the **Table 4** indicate that only Age had a significant effect on task performance, with older participants showing better self-voice recognition. This effect might be related to the performance improvement observed with longer time after last hospitalization, suggesting better clinical remission or slower disease progression in these patients.

**Table 4**. Linear-mixed effects model results: SOVD task accuracy with age and gender effects for all participants.

| Effect | Estimate | t value | p value |
|---|---|---|---|
| (Intercept) | 0.451 | 4.023 | 0.000133 *** |
| Group: AVH- | -0.031 | -0.452 | 0.652546 |
| Group: AVH+ | -0.074 | -1.192 | 0.236354 |
| Voice: Self | -0.131 | -8.388 | p < 0.001 *** |
| Condition: Bone | -0.069 | -4.455 | p < 0.001 *** |
| Gender: M | -0.021 | -0.411 | 0.682034 |
| Age | 0.006 | 2.059 | 0.042920 * |
| Group: AVH- x Voice: Self | 0.037 | 1.315 | 0.188542 |
| Group: AVH+ x Voice: Self | 0.049 | 1.897 | 0.057833 . |
| Group: AVH- x Condition: Bone | 0.065 | 2.331 | 0.019756 * |
| Group: AVH+ x Condition: Bone | 0.064 | 2.498 | 0.012514 * |
| Voice: Self x Condition: Bone | 0.061 | 2.781 | 0.005427 ** |
| Group: AVH- x Voice: Self x Condition: Bone | -0.082 | -2.078 | 0.037772 * |
| Group: AVH+ x Voice: Self x Condition: Bone | -0.059 | -1.631 | 0.102827 |
